# Ketamine Sedation Facilitates Asleep DBS: a multicenter retrospective study

**DOI:** 10.1101/2021.12.12.21267680

**Authors:** Halen Baker Erdman, Evgeniya Kornilov, Eilat Kahana, Omer Zarchi, Johnathan Reiner, Achinoam Socher, Ido Strauss, Zvi Israel, Hagai Bergman, Idit Tamir

**Affiliations:** Department of Medical Neurobiology, Hebrew University, Jerusalem, Israel; Department of Anesthesiology, Rabin Medical Center, Beilinson Hospital, Petach Tikvah, Israel; Department of Neurobiology, Weizmann Institute of Science, Rehovot, Israel; Intraoperative Neurophysiology Unit, Rabin Medical Center, Beilinson Hospital, Petach Tikvah, Israel; Department of Neurology, Rabin Medical Center, Beilinson Hospital, Petach Tikvah, Israel; Department of Neurology, Tel Aviv Sourasky Medical Center, Tel Aviv, Israel; Sackler School of Medicine, Tel Aviv University, Tel Aviv, Israel; Department of Neurosurgery, Tel Aviv Sourasky Medical Center, Tel Aviv, Israel; Department of Neurosurgery, Hadassah Medical Center, Hebrew University, Jerusalem, Israel; The Edmond and Lily Safra Center for Brain Sciences, Hebrew University, Jerusalem, Israel; Department of Neurosurgery, Rabin Medical Center, Beilinson Hospital, Petach Tikvah, Israel

**Keywords:** Deep brain stimulation, Parkinson’s disease, electrophysiology, beta oscillations, ketamine

## Abstract

Deep brain stimulation (DBS) is commonly and safely performed for selective Parkinson’s disease patients. Many centers perform DBS lead positioning exclusively under local anesthesia, to allow for brain microelectrode recordings (MER) and testing of stimulation-related therapeutic and side effects. These measures enable physiological identification of the DBS targets based on electrophysiological properties like firing rates and patterns, optimization of lead placement accuracy, and intra-operative evaluation of therapeutic window. Nevertheless, due to the challenges of awake surgery, some centers use sedation or general anesthesia, despite the distortion of discharge properties, and potential impact on clinical outcomes. Thus, there is a need for a novel anesthesia regimen that enables sedation without compromising intra-operative monitoring. This study investigates the use of low-dose ketamine for conscious sedation during lead positioning in subthalamic nucleus (STN) DBS for Parkinson’s disease patients.

Three anesthetic regimens were retrospectively compared in 38 surgeries across three DBS centers: 1) Interleaved propofol-ketamine (PK), 2) Interleaved propofol-awake (PA), and 3) Fully awake (AA).

All anesthesia regimens achieved satisfactory MER. Automatic detection of STN borders and subdomains using a Hidden Markov Model was similar between the groups. Patients’ symptoms and cooperation during stimulation testing in the ketamine group was not altered. No major adverse effects were reported in the different anesthesia protocols.

These results support the use of low-dose ketamine as a novel alternative for the existing DBS anesthesia regimens, optimizing patient’s experience while preserving lead placement accuracy. A prospective study should be performed to confirm these findings.

## Introduction

Deep brain stimulation (DBS) surgery targeting the subthalamic nucleus (STN) is indicated for patients with advanced idiopathic Parkinson’s disease^1–4^. The surgery is usually carried out with the patient’s head fixated with a stereotactic frame or head pins, and involves three major stages: First, a skin incision and burr hole in the skull over the desired entry point is made; Second, a recording electrode is lowered towards the target, microelectrode recording (MER) is performed to better localize the target nucleus and its borders, high-frequency stimulation is then performed to evaluate the therapeutic window; Third, the recording electrode is removed and the lead is implanted and fixated to the skull, skin closure is performed and frame/pins are removed from the patient’s head. This procedure is carried out for either one or both hemispheres. Subcutaneous tunneling and connection of the lead extenders to an internal pulse generator (IPG) implanted on the patient’s chest or abdomen are performed either as a continuation of the same surgery or in a separate surgery, under general anesthesia.

The procedure details and flow vary among medical centers. For example, not all centers use microelectrode recording to optimize the localization of the DBS leads. These centers rely on anatomy only, using pre- and/or intra-operative neuroimaging scans. However, anatomical subdomains and borders are challenging to define with standard MRI/CT. This approach shortens the surgery but potentially could result in electrode misplacement due to sub-optimal target border detection, stereotactic errors and/or brain shift, which might affect clinical outcome^5–10^.

The anesthetic regimen used for DBS surgery also varies between centers, from awake with local anesthesia only, to monitored anesthesia care with mild sedation, to general anesthesia. Similarly, different drugs are used by different centers. However, propofol-based sedation is typically preferred for the first and third parts of the lead placement (scalp and skull opening and closure and frame removal), due to its rapid pharmacokinetics. MER is traditionally done awake, as propofol and other commonly used sedatives significantly alter the recorded brain signal, the patient’s symptoms, and the ability of the patient to cooperate with the neurological examination during the stimulation testing. The major challenge for the anesthesiologist during surgery is to provide the surgeon with optimal operative conditions (mainly blood pressure management, patient cooperation, and non-altered electrophysiology), while assuring no anesthetic complications (e.g., sedation-induced apnea, increased blood pressure) and non-traumatic patient experience.

Often, the sedation protocol determines the ability to use MER and conduct stimulation testing, depending on the arousal and cooperation level of the patient under the sedative in use. The STN has a very distinctive MER signature which makes recordings useful for successful lead placement. The STN’s characteristic spike shape, discharge rate and pattern (e.g., bursting activity) can be observed in the high pass filtered (spiking) data. Real-time analysis of this data can also be used to aid STN identification. Two of the main landmarks extracted from STN MER are 1) the increase in the total power of spiking activity as measured by the normalized root mean square (RMS) at the entrance to the STN and 2) the increase in power of the Beta frequency band in the dorsolateral oscillatory region (DLOR), which comprise the motor subdomain of the STN^11–13^. Combined, these signatures allow precise localization and placement of the DBS lead contacts to assure optimal surgical outcome.

Various sedative and anesthetic drugs have been shown to distort the typical MER signature of the STN, putting in favor fully awake anesthetic protocol^14–16^. Nevertheless, it is not uncommon for awake DBS cases to be altered or aborted due to marked anxiety and non-cooperative patient responses, leading to sub-optimal surgical results in terms of lead accuracy, clinical outcome, patient’s experience, and other surgical complications^17^.

Ketamine, an *N*-methyl-D-aspartate (NMDA) antagonist, is a dissociative anesthetic discovered over 60 years ago, and has been in wide clinical use ever since. Nowadays it is commonly used in pre-hospital treatment of trauma patients, despite past arguments citing increasing intra-cranial pressure and pro-epileptic effects^18–21^. In recent years, low dose ketamine has also been used for the treatment of major depressive disorder, and its anxiolytic, cognition enhancement, and anti-epileptic effects are now being studied worldwide^22–26^.

Recent animal studies imply that low dose ketamine can be used during MER of the basal ganglia and cortex, without disrupting electrophysiological activity used for DBS navigation^27^. Since both ketamine and propofol have short time constants, the ketamine infusion can be interleaved with propofol sedation periods, as needed. From the anesthesiologist’s perspective, sub-anesthetic ketamine administration might also carry analgesic and anxiolytic advantages during MER, optimizing patient experience and cooperation.

In this study, we propose a novel sedation paradigm for DBS surgeries utilizing the common sedative propofol, a GABA_A_ agonist, during the first and third stages of surgery (scalp and skull opening and closure), and low-dose ketamine during the second (MER) stage. We retrospectively compared the clinical and electrophysiological outcome of Parkinson’s disease patients undergoing STN DBS using either interleaved propofol-ketamine (PK) regimen, interleaved propofol-awake (PA) regimen, or awake-only (AA) regimen.

## Materials and methods

### Participants

A total of 38 patients diagnosed with idiopathic Parkinson’s disease from three medical centers were included in this study. All patients had Unified Parkinson’s disease Rating Scale (UPDRS) part III improvement of >30% in levodopa-challenge test, and underwent DBS surgery targeting the STN. The total number of included MER trajectories and recorded sites were 74 and 5962, respectively.

Three patient groups were defined in this study. 1) Patients that received interleaved propofol-ketamine (PK) sedation, with propofol given during the first and third stages (scalp and skull opening and closure) and ketamine given during the second (MER) stage (12 patients, 23 MER trajectories, 1979 recording sites, Rabin Medical Center). 2) Patients that received an interleaved propofol-awake (PA) sedation, with propofol administered in the first and third surgery stages and no sedatives during the second (MER) stage (5 patients, 11 MER trajectories, 974 recording sites, Tel Aviv Sourasky Medical Center and Rabin Medical Center). 3) Patients that underwent awake surgery (AA) with monitored anesthesia care (21 patients, 40 MER trajectories, 3009 recording sites, Hadassah Medical Center and Rabin Medical Center). Figure 1 outlines the surgical steps and corresponding sedation agent used for each group along with idealized bispectral index (BIS) values^28^.

**Figure 1.**
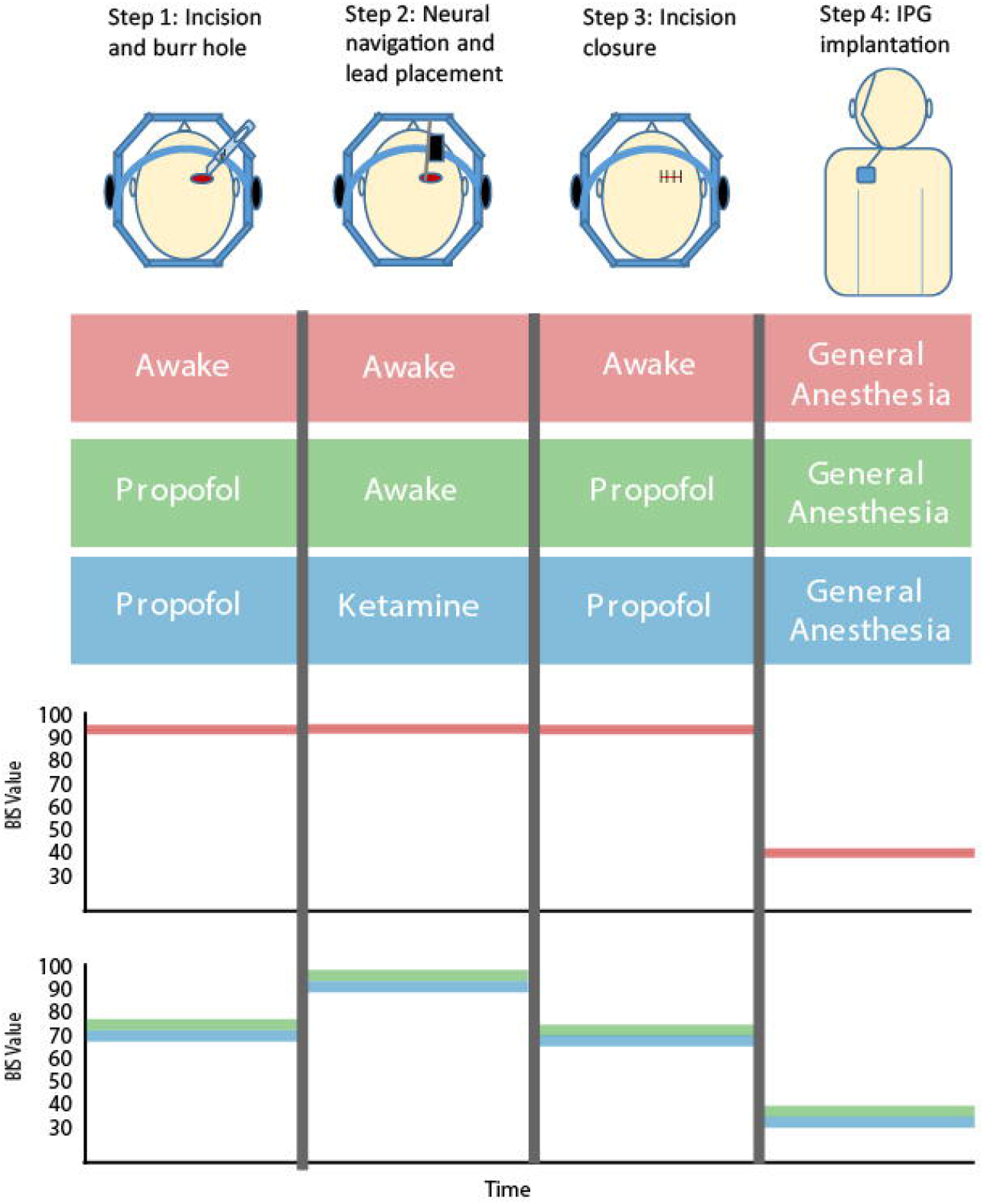
Study’s anesthesia protocols according to DBS surgical stages. **(A)** From left to right: The stages of surgery and corresponding anesthesia given to the experimental group (propofol-ketamine (PK); upper), and the two control groups (propofol-awake (PA; middle) and fully awake (AA; bottom). **(B)** Schematic plot of Bispectral index (BIS) values along DBS procedure in the study groups.

### Study Design

This study is a retrospective study that aims to determine the effectiveness and safety of ketamine administration in DBS surgeries. Informed consent was waived due to the retrospective nature of the study. The study was approved by the hospitals’ Helsinki committee (ethics numbers 0569-20: Rabin Medical Center, 0168-10-HMO: Hadassah Medical Center, and 0092-21-TLV: Tel Aviv Sourasky Medical Center).

### Surgery protocol

DBS electrodes were implanted, targeting the STN as previously described^29^ with the second lead contact at the STN DLOR ventral border, as defined by MER (Figure 2). Briefly, the STN was directly identified and targeted on a pre-operative axial 1.5T or 3T T2-weighted magnetic resonance image (MRI) as a signal hypo-intensity, lateral to the anterior border of the red nucleus and superior to the lateral part of the substantia nigra. The trajectory for lead insertion was chosen based on 3D gadolinium enhanced T1MRI sequences, utilizing the trans-frontal approach, and avoiding eloquent brain areas, blood vessels, and ventricles. On the morning of surgery, a stereotactic frame (either CRW (Integra Lifesciences, Princeton, NJ) or Leksell (Elekta, Crawley, UK)) was fixated on the patient’s head under local anesthesia infiltration (lidocaine 2% and bupivacaine 0.5%, ∼20 cc) and mild sedation (2 mg IV midazolam). A head Computed Tomography (CT) was then performed with a fiducial box attached to the stereotactic frame. CT to MRI fusion was then conducted and frame coordinates were extracted from the navigation system (Medtronic Stealth 8 or Brainlab iPlanNet version 3). The patient was transferred to the operating room and put in a supine position, and the frame was fixed to the operating table. Cefazoline 2 gr IV or clindamycin 900 mg IV was given as a preoperative antibiotic prophylaxis.

**Figure 2.**
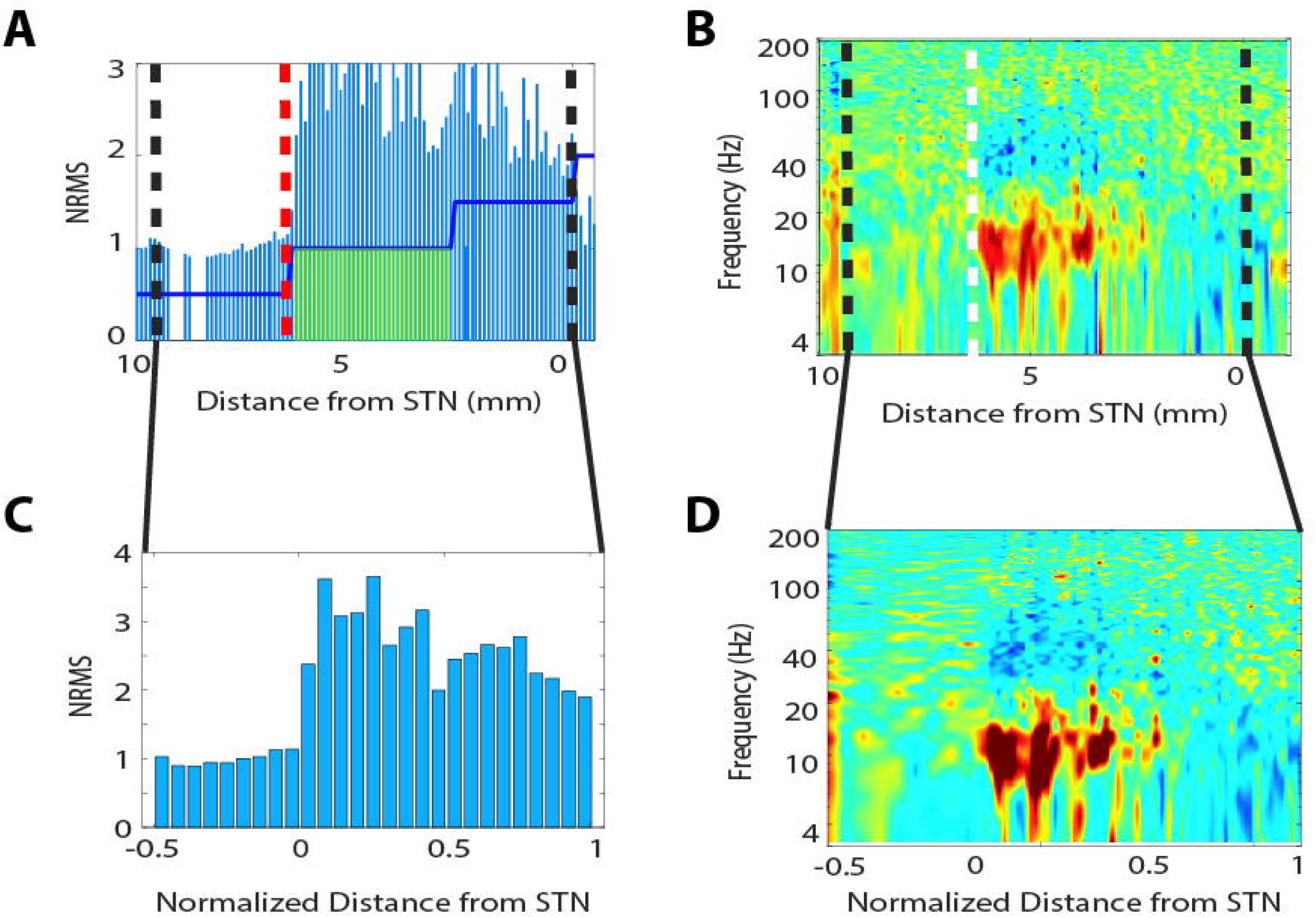
Three-dimensional reconstruction of implanted DBS leads from an example patient. Axial view and close-up of reconstructed leads on Montreal Neurological Institute (MNI) brain from LeadDBS. Example patient was taken from the PK group. Red nucleus is shown in red, STN is shown in orange.

Patients were monitored according to the American Society of Anesthesiologists (ASA) guidelines (electrocardiogram, Sp02, non-invasive or/and invasive blood pressure and EtCO2 nasal monitoring). Level of sedation was assessed clinically. In a few cases, bispectral index (BIS™; Medtronic, Minneapolis, MN) monitor was attached.

In the PA and PK groups, continuous infusion or a bolus of propofol (see dosage in Table 1) was started in preparation for the skin incision and drilling of the burr hole and used to maintain the patient at a moderate level of sedation. Propofol infusion was stopped when the burr-hole drilling ended, 5-20 minutes before MER started. The PK group was then given low dose ketamine either by bolus and/or continuous infusion (see dosage in Table 1). The lead fixation device was fixated to the skull, the dura, arachnoid and pia were coagulated and opened sharply and 1-2 guide tubes were lowered 15-25 mm above the planned target. NeuroProbe microelectrodes (Alpha Omega Engineering, Ziporit, Israel) were then lowered through the guide tubes to 10 mm above the planned target. MER identification of the STN was performed by an expert electrophysiologist, aided by Hidden Markov Model (HMM) STN detection program^11,29–31^ (HaGuide, Alpha Omega Engineering, Ziporit, Israel) using delineation of STN borders and subdomains. Monopolar cathodal stimulation through the macro-contact of the NeuroProbe electrode (130 Hz, 60 μS pulse width, 0.25-5mA) was applied at the lower border of the oscillatory (motor, DLOR) domain of the defined target, to evaluate the therapeutic window. If the therapeutic window was found to be acceptable, the recording electrodes were replaced by a permanent DBS lead (either 3389, Medtronic, Minneapolis, MN or Vercise, Boston Scientific, Marlborough, MA) with the second contact from the tip at the stimulation site. The leads were then secured through the electrodes fixation device, the wound was closed, and the frame was removed from the patient’s head.

**Table 1.** Patient demographics and perioperative characteristics.

|  | **PK, N = 12** | **PA, N = 5** | **AA, N = 21** | p-value |
| --- | --- | --- | --- | --- |
| **Patients' demographics** |  |  |  |  |
| Age, years | 67.5 (58.5-73) | 64 (58.5-67) | 59 (55-65.25) | 0.19 |
| Weight, kg | 73 (61.5-86.5) | 80 (64.5-80) | 66 (60-80) | 0.74 |
| Male sex, % | 9 (75%) | 4 (80%) | 14 (66.7%) | 0.79 (Chi-square) |
| **Preoperative scores** |  |  |  |  |
| Disease duration, years | 8 (4.5-10.5) | 7 (5-9.75) | 8 (7-11) (n = 20) | 0.6 |
| Daily levodopa equivalent, mg | 970.5 (666.5-1445) | 1080 (748.25-1155) | 880 (800-1161.25) (n = 19) | 0.96 |
| *Motor scores* |  |  |  |  |
| UPDRS-III off medication | 41 (32.5-53) (n = 11) | 41.5 (36.5-49) (n = 4) | 40 (29.75-44.5) (n = 17) | 0.64 |
| UPDRS-III on medication | 25 (15.5-30.75) (n = 11) | 21 (15.5-27.75) | 19 (9-27) (n = 18) | 0.47 |
| *Cognitive scores* |  |  |  |  |
| MMSE | 28 (27-29) (n = 10) | 28 (27.5-29) | 29 (27-29) (n = 10) | 0.51 |
| Montreal Cognitive Assessment | 25 (23-27.75) (n = 11) | 26 (23.75-26.75) (n = 3) | 22 (19.75-25.75) (n = 3) | 0.44 |
| Beck's Anxiety Inventory | 13.5 (7-17) (n = 10) | 5 (5-14) (n = 3) | 25 (25-25) (n = 1) | 0.22 |
| Beck's Depression Inventory | 13 (9-21) (n = 10) | 10 (7-24.5) (n = 4) | 13 (8.5-18.5) (n = 11) | 0.94 |
| **Intraoperative electrophysiology** |  |  |  |  |
| Trajectory duration, min | 17 (15-20)* p < 0.001 | 19 (17.5-21)* p < 0.001 | 13 (12-14) | < 0.001 |
| **Anesthesia drugs dosage** |  |  |  |  |
| Propofol total dose per surgery, mg | 565 (391-765) | 299 (244.25-492.5) | NR | 0.052<sup>#</sup> |
| Propofol drip rate, mg/kg/hr | 3.15 (2.84-4.41) | 1.07 (0.42-2.19) | NR | 0.004<sup>#</sup> |
| Ketamine drip rate, mg/kg/hr | 0.12 (0.1-0.21) (n = 8) | NR | NR | NR |
| Total ketamine dose given in bolus, mg | 15 (8-30) (n = 10) | NR | NR | NR |
| Total ketamine dose per surgery, mg | 36 (23-68.5) | NR | NR | NR |
| **Intraoperative antihypertensive drugs** |  |  |  |  |
| Labetalol, mg | 20 (1.25-104)* p = 0.03 | 20.2 (15-57.5) | 0 (0-15) | 0.017 |
| Isosorbide dinitrate, mg | 0 (0-4)* p = 0.02 | 0 (0-0) | 0 (0-0) | 0.017 |
| Hydralazine, mg | 0 (0-5) | 0 (0-2) | 0 (0-0) | 0.191 |
| Nicardipine, mg | 0 (0-0) * p = 0.006 | 0 (0-0) | 0.5 (0-2.5) | 0.003 |

Data presented as median (interquartile range). P-value was calculated with the Kruskal-Wallis test, with Bonferroni post-hoc comparisons.
PK = propofol-ketamine group, PA = propofol-awake group, AA = awake-awake group, UPDRS-III = Unified Parkinson's disease rating scale, Motor Examination. MMSE = Mini-Mental State Examination. IPG = internal pulse generator. NR = not relevant. N = total number of patients in the group. n = number of patients had the parameter measured.
\*significantly different from AA group
<sup>#</sup> Mann-Whitney test.

Some patients underwent a unilateral procedure, while most underwent a bilateral one. In bilateral DBS cases, skin incision, burr-hole drilling and MER localization of the STN of the surgery either took place one after the other for the first side and then for the contralateral side, or skin incision and burr-hole drilling was first conducted for both sides followed by MER for both sides.

Most patients underwent lead extenders and pulse generator implantation (either Activa PC (Medtronic, Minneapolis, MN) or Vercise PC (Boston Scientific, Marlborough, MA)) following lead implantation (same day procedure), while others had it done on a different day. In all patients, this was performed under general anesthesia and tracheal intubation. Following surgery completion, patients were extubated and transferred to the post-anesthesia care unit (PACU).

### Anesthesia Protocols for DBS lead placement

The anesthesia protocols are described in Figure 1.

AA: The fully awake group was given only local anesthetic as a sub-cutaneous injection at the incision sites.

PA: The propofol-awake group was given intravenous propofol for moderate sedation during the incision and burr hole drilling (stage 1), as well as after MER stage for wound closure (stage 3). During MER (stage 2) the patients were kept awake, without any drug administration.

PK: The propofol-ketamine patients were moderately sedated with propofol during the incision and burr-hole drilling (stage 1) and switched to sub-anesthetic ketamine dose during MER (stage 2). Following completion of MER and stimulation testing, ketamine was stopped and propofol was resumed for wound closure and frame removal (stage 3, see Figure 1).

Intraoperative blood pressure was maintained within normal limits (mean arterial pressure 65-85 mmHg, systolic blood pressure < 150 mmHg) and in cases of hypertension was controlled with labetalol, isosorbide dinitrate, hydralazine or nicardipine (Table 1), according to center preferences.

### Electrophysiological data collection

For each hemisphere, one or two microelectrodes were lowered along the planned trajectory using the BenGun electrode holder (NeuroOmega navigation system, Alpha Omega Engineering. Ziporit, Israel). Microelectrode recording began 10 mm above the planned target and proceeded in steps of 0.4 mm prior to STN entrance and in steps of 0.1 mm within the STN. STN borders and subdomains were defined according to electrophysiology findings. Recording at each site began after a 2 second stabilization period and lasted 4 seconds, for a total of 6 seconds at each site.

### Electrophysiological data analysis

Spiking data (300-6000 Hz band-pass filtered) was extracted from the NeuroOmega navigation system (Alpha Omega Engineering, Ziporit, Israel). STN borders and subdomains were determined using a previously published algorithm^11,12^ and verified by an expert electrophysiologist. Briefly, power spectrum density (PSD) and root mean square (RMS) were calculated for each recording site. The RMS is generally reflective of overall neural activity at a certain site and is normalized to the RMS of the first 5-6 sites (usually while the electrode is still in white matter of the internal capsule). The PSD is normalized to the RMS and gives detailed information about the distribution of power at discrete frequency domains (1/3 Hz resolution) as a fraction of the total power at a site.

The recorded STN length was extracted from trajectory recordings for each patient and the length was normalized to a length of 1. For each trajectory, a segment before STN entrance that is equal to half of the length of the STN from that trajectory is included in our normalized visualizations. Figure 3 illustrates this process on the data of one patient. The percent of STN length in which beta oscillations were detected (% DLOR) was analyzed. The amount of beta power within the STN compared to the amount of beta power outside the STN was calculated as the beta ratio for each trajectory and then analyzed across groups. The peak value and area under the curve of the normalized RMS was computed for each trajectory and then evaluated across groups. PSD and normalized RMS (NRMS) for the STN of individual patients were then grouped and averaged to create an average PSD and RMS for each patient group. All analysis was performed using custom Matlab (v2017a) scripts.

**Figure 3.**
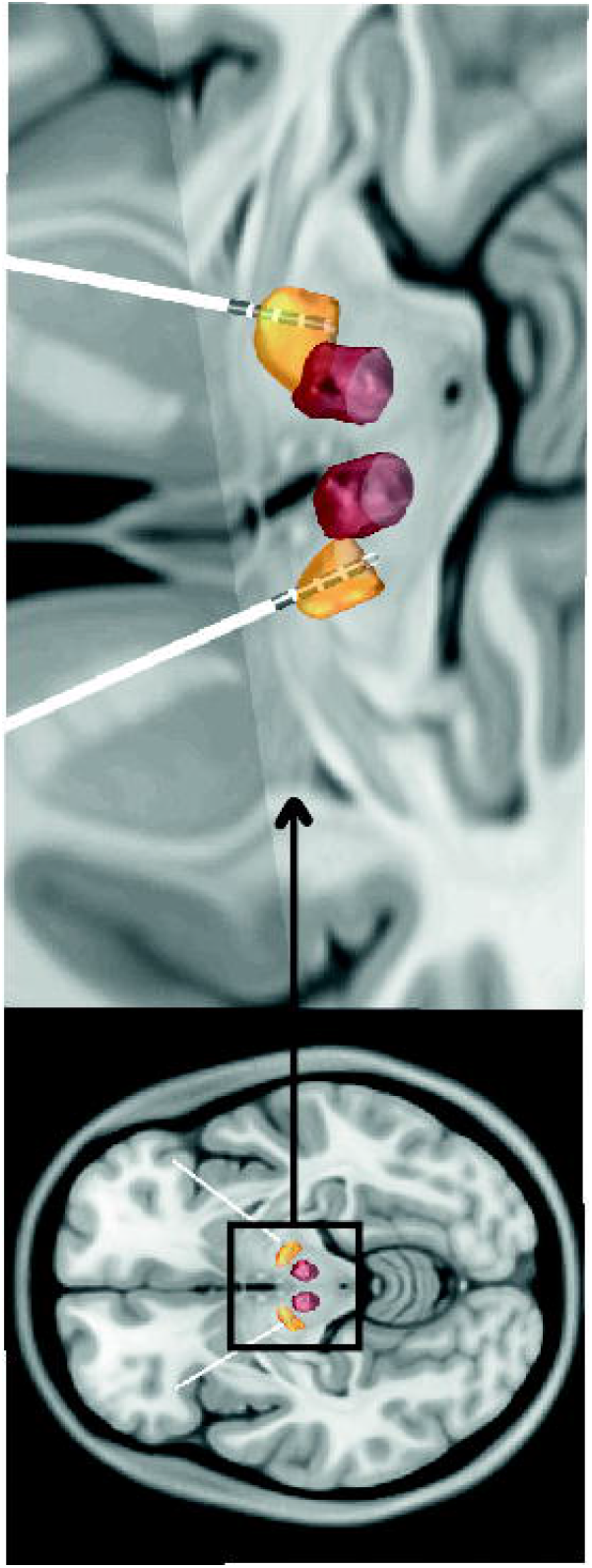
Exemplary STN activity under Ketamine sedation. **(A)** RMS of an example study patient from the PK group, as a function of the estimated distance from target. Black dashed lines indicate the data used for normalization of distance by the STN length). Hidden Markov Model (HMM) results are depicted by the blue solid line. The steps represent (from left to right): 1. White matter area recorded prior to STN entrance; 2. Dorsolateral oscillatory region (DLOR) of the STN (also represented by the translucent green block and entrance indicated by red dashed line); 3. Ventromedial non-oscillatory region (VMNR) of the STN; 4. Exit from STN. **(B)** PSD of the same patient. The dashed white line represents the entrance to the STN. Black dashed lines indicate the data used for normalization (Figure 2D). **(C)** RMS from between black dashed lines of panel A with respect to the normalized length of the STN (0=entrance, 1=exit). **(D)** PSD from between black dashed lines of panel B with respect to normalized STN length.

### Statistical analysis

After the preparation of the database, statistical assumptions were verified. In case of normal distribution (electrophysiological data parameters), parametric tests were used, while nonparametric tests were used for non-normally distributed data (demographic patient data). Results were presented as mean ± standard error mean (SEM) for continuous variables, median (interquartile range) for non-normally distributed data, and number (%) for categorical variables. Analysis of variance (ANOVA) or Kruskal-Wallis test with post-hoc Bonferroni correction were used for group comparisons. For categorical data comparisons, Chi-squared test was performed. Significance was determined using a p-value < 0.05.

### Data availability

Anonymized data available upon request from the corresponding author.

## Results

### Patient and surgery characteristics are similar between groups

Patient demographics and peri-operative characteristics were compared between the three anesthesia protocol groups (AA, PA, and PK). These are detailed in Table 1. Patient demographics and disease – related clinical characteristics including age, disease duration, pre-operative disease severity (motor symptoms, characterized by Unified Parkinson’s Disease Rating Scale; UPDRS III) and medication dose prior to surgery were similar between the three anesthesia regimen groups. Surgical and anesthesia characteristics also did not differ between the groups, except for mean trajectory time with the PK and PA groups averaging 18 and 19 minutes, respectively, and the AA group averaging 13 minutes per trajectory. These differences probably reflect the non-even distribution of patients between the centers and the different DBS routines.

### Anatomical and electrophysiological target characteristics are similar between groups

Intra-operatively, the STN could be recognized electrophysiologically in all MER trajectories. Mean recorded STN length, mean DLOR length and mean fraction of DLOR out of STN length (% DLOR) were similar between the groups (p = 0.08, p= 0.76, and p = 0.69, respectively) (Figure 4A-C).

**Figure 4.**
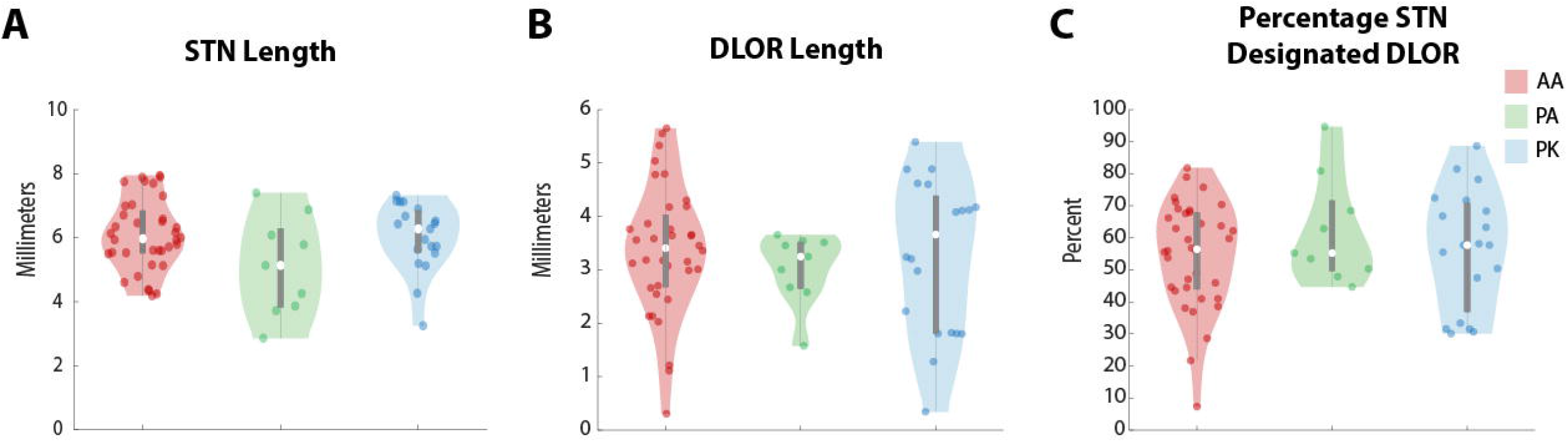
Comparison of STN characteristics across study groups. AA group is indicated in red, PA group is indicated in green and PK group is indicated in blue. **(A)** Shows the median STN length, **(B)** shows the median DLOR length and **(C)** shows the median percentage that was designated DLOR. Median value is denoted by a white circle. The dark grey bar indicates the 25^th^ to 75^th^ percentile. The whiskers extend to the maximum and minimum values, excluding outliers. The coloured translucent shape encompasses all data points, including outliers, where the width is a function of data density.

Root mean square analysis revealed a similar pattern for all three groups (Figure 5, upper panels). Namely, a ramped increase from 1 to ∼2.5-3 NRMS at the entrance to the STN and a return to baseline at the exit from the STN was observed. Example spiking data from before STN entrance and within the STN are shown in Figure 5B,C from one patient per group. Peak STN NRMS values were 4.57 ± 0.40 for the PK group, 3.99 ± 0.29 for the PA group and 5.10 ± 0.26 for the AA group (p = 0.15) (Figure 6A). The total area under the curve for the PK group was 146.9 ± 11.83 compared to 137.4 ± 12.06 and 148.8 ± 7.19 in the PA and AA groups, respectively (p = 0.81; Figure 6B).

**Figure 5.**
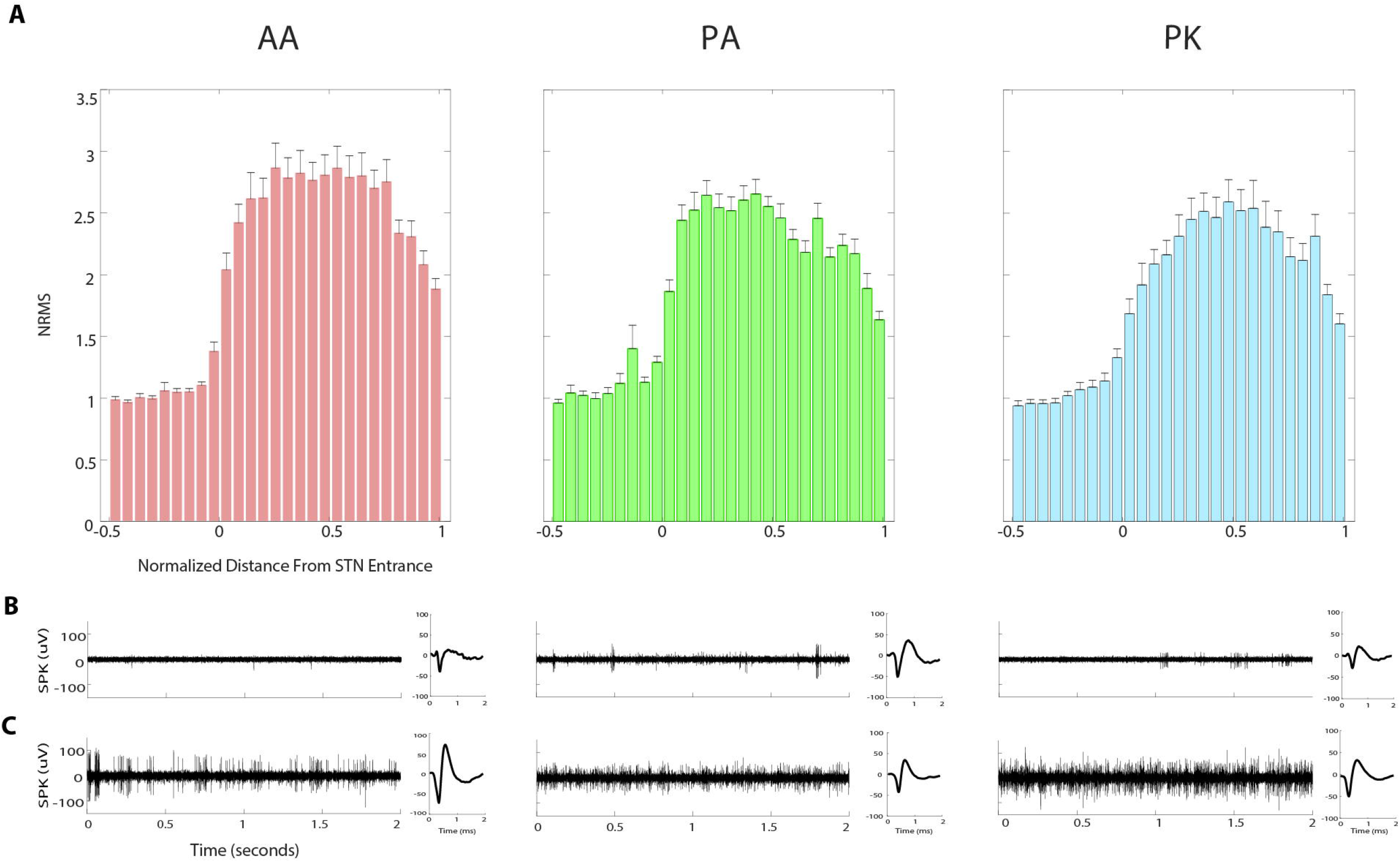
Comparison of spike and RMS activity between study groups. **(A)** RMS of the AA group (red), the PA group (green) and the PK group (blue) along the recorded STN trajectory. All groups show a sharp increase in activity at the entrance to the STN (marked 0) and sharp decrease at the exit from the STN (marked 1). **(B)** Corresponding examples of spiking activity (SPK) data traces prior to STN entrance in the respective groups and average spike shape of the data shown. **(C)** Corresponding examples of spiking activity (SPK) data traces in the motor STN region of the respective groups and average spike shape of the data shown.

**Figure 6.**
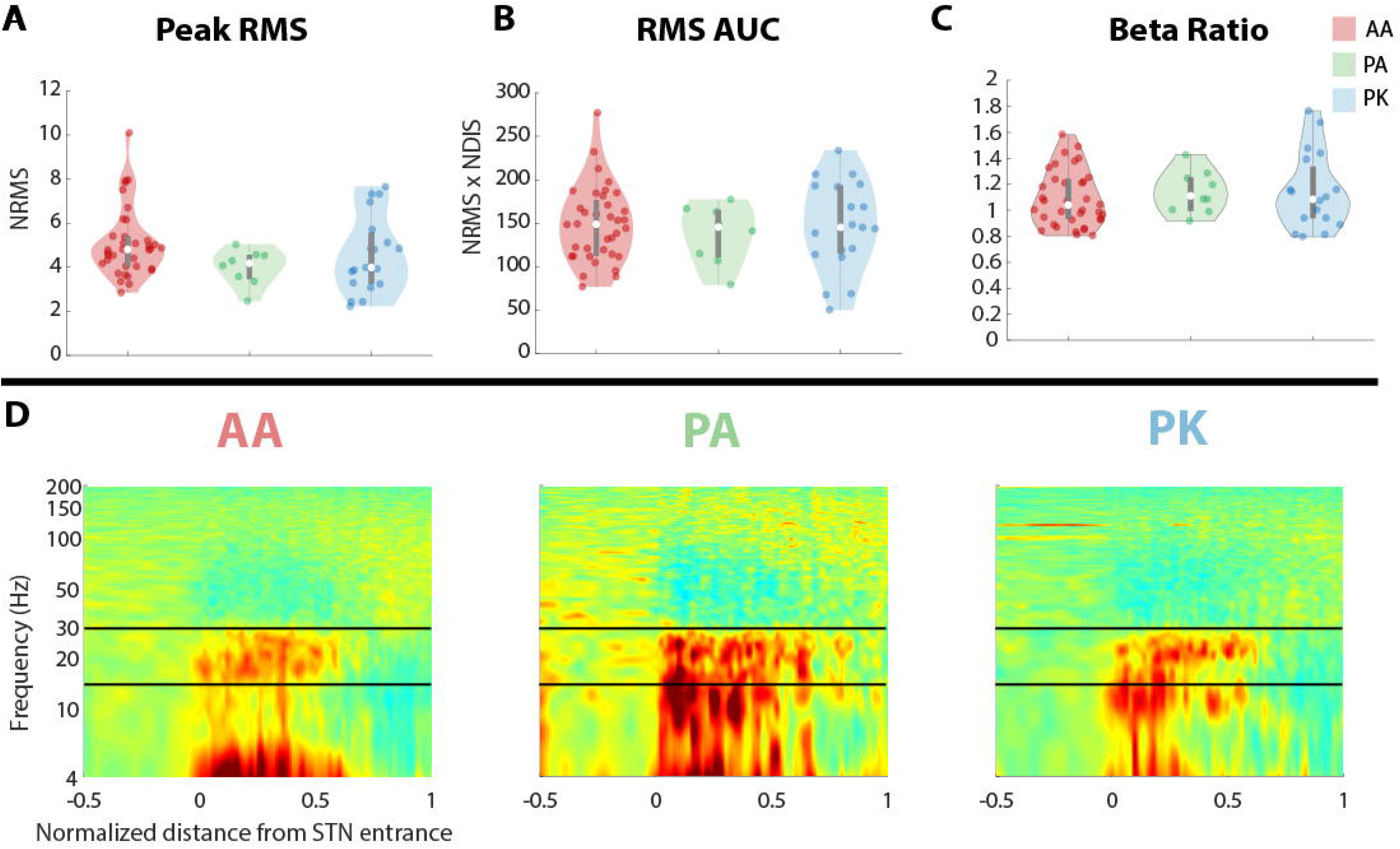
Electrophysiological Characteristics and Power Spectrum Density (PSD) across study groups. AA group is indicated in red, PA group is indicated in green and PK group is indicated in blue. **(A)** The median peak normalized RMS value. **(B)** The median area under the curve (AUC) of the RMS. **(C)** The median beta ratio calculated for each group. **(D)** PSD of the AA group, the PA group, and the PK group. STN motor region entrance is indicated by increased power in the beta frequency range (denoted by solid black lines). Structure of **(A-C)** are as in Figure 4.

Spectral analysis of the spiking data again revealed a similar pattern across the groups (Figure 6D). Consistent with previously published literature^32–34^, the entrance of the STN was marked by a shift in power to beta frequency extending throughout the DLOR. At the transition from the DLOR to the ventro-medial non oscillatory region (VMNR, non-motor) sub-section of the STN, power was again dispersed. The relative beta power in the STN compared to outside the STN (beta ratio) did not vary between the groups (*p* = 0.63; Figure 6C).

### Operative outcome and patient experience

No significant intra- or post-operative adverse effects were reported in the three anesthetic regimens. All patients completed surgery as planned. No hemorrhage, infection, death, or lead misplacement needing surgical revision was reported in the three groups. None of the patients had intra- or post-operative seizure. All three groups showed similar levels of cooperation and arousal during the stimulation and therapeutic window assessment stage. Only one patient in the ketamine group reported intra-operatively of visual hallucinations. However, these did not interfere with the surgical workflow and the patient did not report them as a “negative” experience. Blood pressure was well controlled in all patient groups. (Table 1).

## Discussion

In this study we compared the use of an interleaved propofol-ketamine anesthesia regimen to interleaved propofol-awake or awake only anesthetic regimens during DBS surgery for Parkinson’s disease, targeting the STN. The results of this study suggest a novel ketamine-based regimen for conscious anesthesia in patients undergoing DBS surgery. It shows the safety of using an interleaved propofol-ketamine sedation in DBS cases is not inferior to other anesthesia regimens in which patients are awake throughout recordings and stimulation. MER quality under ketamine is shown here to be comparable with MER performed completely awake, without using any systemic anesthetics prior to or during recordings. Moreover, it proves the ability to use a potent anesthetic agent during DBS surgery, without compromising intra-operative MER, clinical stimulation testing and post-operative clinical outcome. Due to its retrospective nature, this study is limited in its ability to show the effectiveness of the ketamine regimen in improving patients’ overall surgical experience and surgical outcome. Nevertheless, such improvement may result due to the ketamine’s immediate and short-lasting anxiolytic and dissociative effects. We therefore believe that ketamine anesthesia might bridge the gap between optimal patient comfort and accurate electrode placement.

### The use of ketamine in neurosurgery

Since its introduction in 1970, ketamine has grown to have a bad reputation due to concerns that it may cause an increase in intracranial pressure, have pro-epileptic effects, possible frightening hallucinations and abuse potential. Despite many papers dispelling these worries^22,25,35,36^, there still seems to be some concern among anesthesiologists. The present study not only adds to the evidence that low dose (sub-anesthetic) ketamine can be safely used, but in fact highlights its advantages compared to other sedative hypnotic drugs. Ketamine has been reported to attenuate post-operative cognitive dysfunction, improve postoperative analgesia and reduce inflammatory responses during surgery^22,35,37^. In contrast to classic sedatives like GABA-receptors agonists, ketamine is a dissociative drug that does not suppress cardio-pulmonary homeostatic reflexes and does not mask brain electrophysiological activity^27^.

Ketamine’s use in DBS surgeries has not been well studied. One report suggested the use of ketamine in DBS surgeries for patients with severe tremor and dyskinesia but did not discuss the effect on MER and reported higher doses than were used in this study^38^. Other studies on DBS in pediatric population have used ketamine in combination with other sedatives and have reported that ketamine does not significantly affect MER^39,40^. Ketamine has been effectively used in other types of surgeries and seems to not only be well tolerated but also to provide a host of post-operative benefits^22,26,35,41,42^.

### Anxiolytic effects of ketamine

Besides the demonstrated ability to perform accurate electrophysiological recordings, sub-anesthetic ketamine sedation also allows for intraoperative interaction with the patient. This is extremely important during the therapeutic window assessment and identification of stimulation-induced therapeutic and side effects. A small therapeutic window might intra-operatively hint at a sub-optimal electrode location, and the nature of the side effects can help in in identifying the direction of the misplacement. This stage of surgery is key to proper electrode placement and to assure straight forward post-operative programming. Low dose ketamine administration produces anxiolysis, while maintaining the patients’ ability to respond and interact with motor and cognitive examination as needed. In fact, the anxiolytic effect of ketamine has led to its use as a treatment for anxiety and depression^24,43–46^.

As a result of the anxiolytic effect of ketamine, some patients might show a decrease in stress-related tremor. However, this effect is usually mild and does not seem to interfere with clinical testing during surgery. Another concern when contemplating any operation on an elderly patient is the effect it may have on the patient’s cognitive state. Evidence suggests that ketamine may actually be neuroprotective^25,26,35,42^. This could be an added benefit for patients already in a fragile state, especially Parkinson’s disease patients approaching surgery with mild to moderate cognitive decline.

Ketamine is a well-known hallucinatory drug^47^, which limits its wide use in anesthesiology. Ketamine induced hallucinations appear in a dose dependent manner^48^. In the patient cohort of this study one patient did experience pleasant hallucinations and described seeing colors and shapes and feeling like they were in a dream-like state, consistent with Jacksonian theory^49^. The nature of the hallucinations might be compounded by the already stressful environment of the operating room. Nevertheless, there is evidence that preceding ketamine with propofol decreases frightening hallucination risk^50,51^. Therefore, we suggest an interleaved propofol-ketamine protocol in order to limit this concern. By restricting ketamine administration to a short time period and monitored low doses, we can decrease the risk of hallucination occurrence. Indeed, in the present study, none of the participants in the PK group reported negative hallucinations during surgery. Current literature suggests that low dose ketamine rarely results in negative hallucinations, even in patients with a history of psychosis^52–55^.

Ketamine’s mechanism of action probably involves the blockade of NMDA receptors on GABAergic neurons in subcortical and limbic regions. This disinhibition leads to an increase in neuronal activity and enhanced dopamine and glutamate release in the cortex and limbic striatal areas^56^. The increase in frontal excitability along with an increase in glutamate binding to AMPA in sensory cortices has been proposed as the cause of the distorted perceptual state following ketamine administration^57^. Sensory evoked thalamic gamma oscillations have also been shown to decrease during ketamine administration, suggesting potential gating along the corticothalamic pathway preventing sensory information from reaching higher areas. Meanwhile, no consensus has been reached as to the mechanism for ketamine’s rapid anti-depression effects, but many studies have focused on the increase in glutamate and resulting neuroplastic changes^58–60^.

Finally, ketamine has sympathomimetic properties that may result in increased blood pressure^61^. Indeed, the patients in the propofol-ketamine group, as well as in the two other groups, received nicardipine, labetalol, isosorbide dinitrate and hydralazine to maintain desired blood pressure levels. However, compared to other neurosurgical procedures, the ideal blood pressure during DBS surgery is on the higher end of normal to avoid expansion of the subdural space which could lead to brain shift and sub-optimal lead positioning. We could not compare antihypertensive drugs use between groups because of different hospital protocols. However, none of the patients had uncontrolled blood pressure or complications related to increased blood pressure during surgery. Furthermore, the increase in blood pressure, when present, was gradual and not rapid, allowing reasonable control with antihypertensive drugs.

In addition to the previously mentioned advantages of ketamine use during MER, the use of ketamine might also be extended to substitute, or to augment, propofol in the first and third stages of surgery. For example, patients that develop snoring, apnea or restlessness under propofol, or are pre-operatively diagnosed with obstructive sleep apnea (which is fairly common in Parkinson’s disease patients^62^) might benefit from having low-dose ketamine as their sole anesthetic for the entire procedure, or in a combination with low doses of propofol.

### Limitations and conclusions

This study used a low (sub-anesthetic) dose of ketamine. Further study is warranted to test the effectiveness and safety of higher doses of ketamine in the setup of DBS. In addition, various administration techniques of ketamine should be studied (bolus vs. continuous infusion). Further, we have used racemic ketamine for this study. However, S-ketamine might be even better suited for DBS as it may facilitate neuroprotective effects^63^. This study used a relatively small number of patients that were gathered over three medical centers, limiting our ability to compare between groups and likely contributed to the difference in trajectory durations between groups. Finally, due to the retrospective nature, no patient questionnaires were used to evaluate overall patient experience and no standardized follow-up was carried out to compare clinical symptom improvement.

Keeping the study limitations in mind, we suggest that ketamine administration can be safely used as an alternative sedation technique for the awake patient during MER and lead localization stage in DBS surgery. It may be especially useful for patients with anxiety regarding the awake phase of DBS surgery, and enhance their level of cooperation. Moreover, its use in the pediatric population undergoing DBS should also be further studied, as well as its possible use in other awake neurosurgical procedures requiring patient cooperation and electrophysiological monitoring like epilepsy surgery and tumor resection surgery. Further prospective research on a larger scale should be performed to validate the findings of this study.

## Data Availability

Anonymized data will be made available upon request from teh corresponding author.

## Abbreviations

AA: awake awake
AMPA: alpha-amino-3-hydroxy-5-methyl-4-isoxazolepropionic acid
ASA: American Society of Anesthesiologists
BIS: bisectral index
DBS: Deep brain stimulation
DLOR: dorsolateral oscillatory region
HMM: Hidden Markov Model
IPG: internal pulse generator
MER: microelectrode recording
NMDA: *N*-methyl-D-aspartate
NRMS: normalized root mean square
PA: propofol-awake
PACU: post-anesthesia care unit
PK: propofol-ketamine
PSD: power spectrum density
RMS: root mean square
SEM: standard error mean
STN: subthalamic nucleus
UPDRS: Unified Parkinson’s disease Rating Scale
VMNR: ventro-medial non oscillatory region

